# Self-determination in minoritized autistic adolescents and adults

**DOI:** 10.1101/2024.05.02.24306799

**Authors:** Teresa Girolamo, Marissa Birmingham, Karina Patel, Samantha Ghali, Iván Campos, Kyle Greene-Pendelton, Rebecca Canale, Caroline Larson, Audra Sterling, Karrie A. Shogren

## Abstract

Self-determination plays an important role in outcomes in autism and shows intersectional disparities. Yet, little is known about the role of individual differences or social drivers of health in the development of self-determination. Understanding these factors is key for developing effective supports. This mixed-methods convergent study examined self-determination in racially and ethnically minoritized autistic individuals and caregivers. Participants ages 13 to 30 (*N* = 73) varying widely in language and cognitive ability and caregivers (*n* =52) completed the Self-Determination Inventory. Autism traits and sense of community predicted caregiver report of self-determination, and autism traits and language predicted self-report of self-determination, consistent with DisCrit and Diversity Science. Self-Determination Inventory interviews of a subset of participants (*n* = 13) and caregivers (*n* = 9) were analyzed using inductive thematic analysis. Themes pointed to the role of the intersection of race and disability in shaping self-determination. Altogether, findings point to the importance of these frameworks, environmental influences, and multi-informant perspectives in characterizing self-determination. Future work should focus on the impact of environmental factors in self-determination in minoritized autistic individuals during the transition to adulthood.

## Self-determination in minoritized autistic adolescents and young adults

In the U.S., the transition to adulthood can be challenging for autistic individuals, who lose access to special education supports and services (*Every Student Succeeds Act* [ESSA], 2015; *Individuals with Disabilities Education Improvement Act of 2004* [IDEIA], 2018). Inadequacy of environmental supports place autistic adults at risk for poor adaptive, educational, occupational, and social outcomes (Billstedt et al., 2010; Kuo et al., 2018). Yet, research tends to overlook environment influences on outcomes (Anderson et al., 2018b), focusing mostly on individual differences in language, cognition, and autism traits (Billstedt et al., 2005; Howlin et al., 2004, 2013; Magiati et al., 2014). To characterize the transition to adulthood, we must also include individual fit within the environment (Lai et al., 2020).

One way to understand environmental fit is self-determination, or sense of ability to set and work toward goals as a causal agent given environmental demands (Shogren et al., 2015). Self-determination emphasizes autonomy and accounts for factors in experiences beyond individual control (Shogren et al., 2020). In adulthood, self-determination impacts myriad domains (Kim, 2019) and is important for post-secondary outcomes (Shogren et al., 2015, 2017a). Assessing self-determination in diverse autistic youth and adults is feasible using the Self-Determination Inventory (SDI; Hagiwara et al., 2021a; Shogren et al., 2018a). However, our understanding is limited. Prior work under-emphasizes environmental influences, within-group heterogeneity (Morán et al., 2021), and experiences of minoritized individuals (Kim, 2019; Thoma et al., 2016). To address these gaps, this report examines self-determination in minoritized autistic adolescents and adults.

### Self-Determination as a Construct

Self-determined people can effect change on the environment to achieve goals (Shogren& Wehmeyer, 2015; Shogren et al., 2018b). Causal Agency explains the process of becoming self-determined, with three essential characteristics that increase self-determination over time (Shogren & Wehmeyer, 2015). Action-control beliefs refer to the sense that an individual feels they can achieve a goal (control expectancies), have the means to achieve a goal (empowerment), and realize those means to effect change in their environment (self-realization; Chang et al., 2017). Volitional actions are self-initiated, aligned with individual preference, and support identification of goals. Agentic actions are self-directed actions that identify pathways to support reaching a goal (Shogren et al., 2017b). In using a social-ecological approach to disability (Bronfenbrenner, 1992; Shogren et al., 2018b), self-determination underlines that multiple levels of environment (interactions, communities) differently provide supports (Bronfenbrenner, 1977).

#### Environmental Influences

Characterizing the environment must consider diversity and social drivers of health (Centers for Disease Control and Prevention, 2021). For instance, complex experiences with racism and ableism impact self-determination in Black students with developmental disabilities (Shogren et al., 2021c; Taylor et al., 2023). Per intersectionality theory (Crenshaw, 1989) and Dis/ability Studies and Critical Race Theory (Annamma et al., 2013), structural racism interacts with other types of discrimination (e.g., ableism) to cause this marginalization. Yet, there is no one-to-one ratio between race, environment, or experiences. This motivates a Diversity Science approach, which focuses on heterogeneity within minoritized communities (Plaut, 2010).

Given these frameworks, examining social drivers of health allows for exploration of heterogeneity (American Psychological Association [APA], 2020). First, while autistic youth lose access to school and child-based services in the transition to adulthood (Eilenberg et al., 2019), individual needs vary (Laxman et al., 2019; Schott et al., 2021; Taylor & Henninger, 2015). Unmet service needs, and not services received, captures this variability (Burke et al., 2023), but it is unclear how unmet service needs impact self-reported self-determination (Cheak-Zamora et al., 2020). Second, barriers to services are higher if caregivers feel a greater burden (Ishler et al., 2023; Taylor & Henninger, 2015); thus, more barriers could be tied to lower caregiver-reported self-determination (Anderson et al., 2018a). Third, caregiver-reported sense of community predicts community participation (Talò et al., 2014), reduces disability-related discrimination, and increases environmental fit (Daley et al., 2018). Sense of community might increase caregiver-reported self-determination. Though important, the impact of these social drivers of health on self-reported self-determination is unknown.

#### Summary

Self-determination is relevant to understanding the experiences of autistic individuals (Thoma et al., 2016). Nuances in self-determination cannot be reduced to disability or race and must include environmental factors, that vary within and between communities.

### Assessment of Self-Determination in Minoritized Autistic Adolescents and Adults

The SDI shows intersectional differences in self-determination that cannot be reduced to race or disability alone (Shogren et al., 2018a, 2021b). The SDI includes 21 items and has been validated across three versions: adolescents (ages 13 to 22) with and without disabilities in the SDI: Student Report (Shogren et al., 2020), adults (ages over 18) with and without disabilities in the SDI: Adult Report (SDI:AR; Shogren et al., 2021a), and with teachers as proxy respondents in the SDI: Parent/Teacher Report (SDI:PTR; Shogren et al., 2021a). All versions are completed online. Respondents make ratings on a slider scale with anchors of “Disagree” and “Agree,” resulting in a computer-generated overall self-determination score of 0 to 99, and scores for volitional action (Decide), agentic action (Act), and action-control beliefs (Believe; Shogren et al., 2015); see Table 1 (Shogren, 2018). Items are written at a third grade reading level, with definitions for complex vocabulary (e.g., “obstacles”) and a read-aloud function (Shogren, 2018).

**Table 1.**
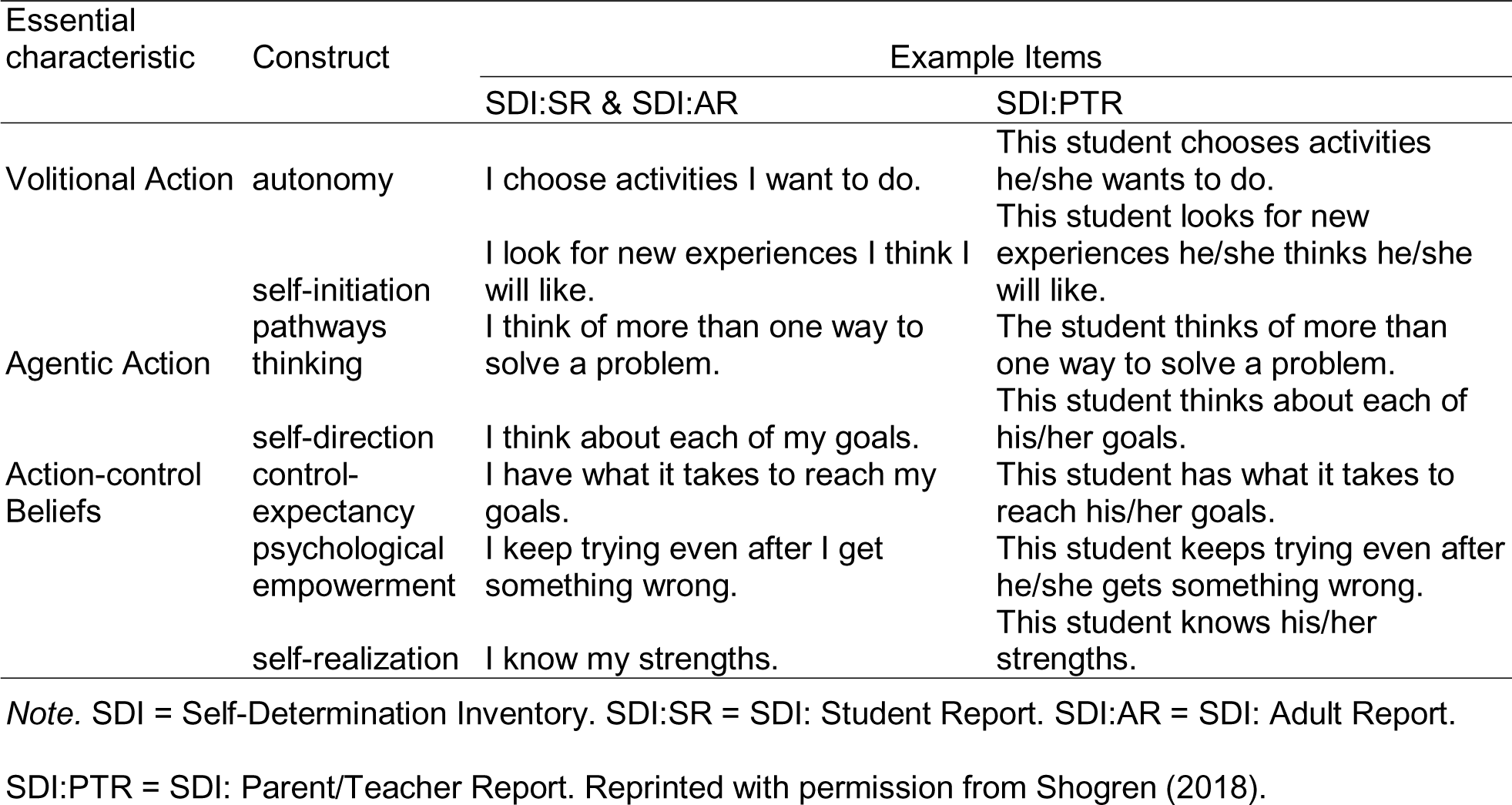
Theoretical Structure of Causal Agency Theory and Example Items of the SDI:SR, SDI:AR, and SDI:PTR.

| Essential characteristic | Construct | Example Items |  |
| --- | --- | --- | --- |
|  |  | SDI:SR & SDI:AR | SDI:PTR |
| Volitional Action | autonomy | I choose activities I want to do. | This student chooses activities he/she wants to do. |
|  | self-initiation | I look for new experiences I think I will like. | This student looks for new experiences he/she thinks he/she will like. |
| Agentic Action | pathways thinking | I think of more than one way to solve a problem. | The student thinks of more than one way to solve a problem. |
|  | self-direction | I think about each of my goals. | This student thinks about each of his/her goals. |
| Action-control Beliefs | control-expectancy | I have what it takes to reach my goals. | This student has what it takes to reach his/her goals. |
|  | psychological empowerment | I keep trying even after I get something wrong. | This student keeps trying even after he/she gets something wrong. |
|  | self-realization | I know my strengths. | This student knows his/her strengths. |

*Note.* SDI = Self-Determination Inventory. SDI:SR = SDI: Student Report. SDI:AR = SDI: Adult Report.
SDI:PTR = SDI: Parent/Teacher Report. Reprinted with permission from Shogren (2018).

#### Intersectional Differences in SDI Scores

SDI scores show nuances, such as lower SDI:SR latent means in diverse with disabilities (α = 70.58-72.99) than white students without disabilities (α = 76.05) (Shogren et al., 2018a) and nonsignificant differences in SDI:AR estimates of autistic adults from adults without disabilities (80.45 versus 84; Hagiwara et al., 2021a). These patterns align with those of other self-determination measures (Chou et al., 2017; Shogren et al., 2006, 2014; Shogren & Shaw, 2017). A limitation in interpreting findings is that studies use primary disability label (Hagiwara et al., 2021a). Though primary label is consistent with service systems, autism often co-occurs with structural language (grammar) impairment (Boucher, 2012) and intellectual disability (Baio et al., 2018), which vary in definition (Girolamo et al., 2023c; Taylor & Henninger, 2015). To build the evidence base, using language, cognitive ability, and autism traits as continuous predictors of self-determination is needed (Kover & Abbeduto, 2023).

Beyond continuous approaches to individual differences, cultural, environmental, and individual influences are important for understanding self-determination. Cultures vary in notions of autonomy (Shogren, 2011), and may be collectivist versus individualistic (Leake & Boone, 2007; Trainor, 2005). There is also sociocultural variation in priorities for goal setting (Zhang et al., 2010). For instance, “I chose what my room looks like” in China may not align with cultural priorities for goal setting (i.e., academics; Xu et al., 2022). Culture also shapes the environment. Lower scores in Spain compared to the U.S. for “I know what I do best” could indicate reduced access to inclusive educational settings or cultural differences (Shogren et al., 2019). Also, cultural norms, lived experiences, and environmental bias influence caregiver perceptions about autism (Dababnah et al., 2018; Rivera-Figueroa et al., 2022). Per DisCrit (Annamma et al., 2013) and Diversity Science (Plaut, 2010), findings point to the importance of understanding nuances beyond scores.

#### Summary

Research supports the utility of the SDI for minoritized autistic individuals but is limited to broad patterns. Understanding self-determination at the intersection of race and disability in greater depth requires considering individual differences, social drivers of health, and individual responses.

### The Current Study

This mixed-methods study focused on self-determination in minoritized autistic adolescents and adults and caregivers. This study used a convergent design (Creswell & Clark, 2017; Pyschology Press, 2013). Aims were:

1) To identify how categorical (language impairment, intellectual disability, levels of autism traits) versus continuous measures of individual differences (language, NVIQ, autism traits) predict SDI self-report and SDI:PTR overall scores, when controlling for social drivers of health (sense of community, unmet service needs, barriers to services);
2) To explore themes in SDI self-report and SDI:PTR interview responses in a subsample of autistic individuals with language impairment and caregivers;
3) To generate integrated support for the relevance of DisCrit and Diversity Science based on quantitative and qualitative data;

Per prior work (Hagiwara et al., 2021a; Shogren et al., 2018a), we expected categorical approaches to individual differences would not predict either score. As continuous measures of language predicted outcomes in autism (Magiati et al., 2014), we expected language to predict SDI self-report and SDI:PTR scores. In both approaches, we expected unmet service needs would predict SDI self-report but not SDI:PTR scores (Cheak-Zamora et al., 2020), and barriers to services and sense of community would predict SDI:PTR scores (Daley et al., 2018; Ishler et al., 2023; Taylor & Henninger, 2015). Per the study design, the second and third aims had no hypotheses.

## Method

This study received institutional board approval. A convergent mixed methods design examined SDI scores and interview themes (Creswell & Clark, 2017; Pyschology Press, 2013). This design allowed for insights into self-determination, given DisCrit (Annamma et al., 2013) and Diversity Science (Plaut, 2010). With little evidence, understanding both scores and themes was important.

### Community Involvement and Researcher Reflexivity

This study used a community-based participatory approach [anonymized]. To support power sharing (Wallerstein & Duran, 2006), partners chose their role and opted to join the team at all study stages. The research team included diverse individuals with lived, personal, and professional experiences with autism, but team members did not know participants. The assumptions were that using a participatory approach to engage participants would be effective, yield meaningful information about self-determination, and provide findings that may not transfer, given systematic exclusion in autism research (Maye et al., 2021).

### Participants

#### Selection Criteria

Selection criteria for autistic individuals were: (a) racially minoritized and/or ethnically minoritized per U.S. Census guidelines (Office of Management and Budget, 1997); (b) formal clinical diagnosis of autism, independently confirmed using the Social Responsiveness Scale-2^nd^ Ed. (SRS-2; Constantino, 2012) and expert clinical judgment; (c) ages 13 to 30, coinciding with approximately when transition planning begins in many U.S. states and 10 years post-federal eligibility for special education services (IDEIA, 2018); (d) proficiency in English per self-report and clinical judgment during screening, as assessments were in English; (e) adequate hearing and vision thresholds for responding to audiovisual stimuli on a computer screen, and; (f) use of spoken language to communicate, as study activities required producing language responses. Selection criteria for caregivers were: (a) caregiving role for autistic participants; (b) proficiency in English; and (c) adequate hearing and vision thresholds for study activities.

#### Recruitment and Procedures

We recruited participants by [anonymized]: (a) sharing study flyers with organizations serving diverse autistic adolescents and adults, (b) providing consultation about the study to individuals and families by phone, Zoom, or email, and (c) obtaining informed consent using a dynamic process. Recruitment and data collection took place from 2022 to 2023 remotely on HIPAA-compliant Zoom, using an embedded design and a stopping rule of 68 for assessment and 17 for interviews (Hennink & Kaiser, 2022). The first author administered a behavioral assessment protocol to participants using test developer guidance (Pearson, 2023). Participants and caregivers completed questionnaires and the SDI as a semi-structured interview. Participants in the first aim were 73 autistic individuals and 52 caregivers; see Table 2. Participants in the second aim were a sub-sample of 13 autistic individuals and 9 caregivers. The sample was primarily male for sex assigned at birth and gender (67-69%; sub-sample: 92%). Most caregivers were mothers (89-90%). Most autistic participants (55%; sub-sample: 100%) scored ≤ −1.25 *SD* on at least two language measures (Tomblin et al., 1997); see Table 3. Six (8.2%) had NVIQ < 70, and 15 (21.1%) had NVIQ of 70 to 84. SRS-2 total *t*-scores varied: high (*n* = 33, or 45.2%), moderate (*n* = 19, or 26.0%), mild (*n* = 10, or 13.7%), and subclinical (*n* = 11, or 15.1%).

**Table 2.** Participant Sociodemographics.

| Variable | All ($N = 73$) | | Subsample ($n=13$) | |
| --- | --- | --- | --- | --- |
|  | *n* | % | *n* | % |
| Chronological age | 19.7 (13.3-30.4) |  | 22.5 (16.5-29.8) |  |
| Race |  |  |  |  |
| American Indian, Native American, or Alaska Native | 12 | 16.4 | 0 | 0 |
| Asian | 19 | 26.0 | 0 | 0 |
| Black | 38 | 52.1 | 10 | 76.9 |
| multiracial | 26 | 35.6 | 1 | 7.7 |
| Pacific Islander | 4 | 5.5 | 0 | 0 |
| white | 20 | 27.4 | 0 | 0 |
| other | 9 | 12.3 | 1 | 7.7 |
| Hispanic/Latine | 13 | 17.8 | 4 | 30.8 |
| Sex Assigned at Birth |  |  |  |  |
| Female | 23 | 31.5 | 1 | 7.7 |
| Male | 50 | 68.5 | 12 | 92.3 |
| Gender |  |  |  |  |
| Female | 24 | 32.9 | 1 | 7.7 |
| Male | 49 | 67.1 | 12 | 92.3 |
| Caregiver |  |  |  |  |
| Grandparent - grandmother | 2 | 3.8 | 0 | 0.0 |
| Parent - mother | 47 | 90.4 | 8 | 88.9 |
| Parent - father | 2 | 3.8 | 0 | 0.0 |
| Sibling | 1 | 1.9 | 1 | 11.1 |

*Note.* Multiracial not mutually exclusive with other racial/ethnic categories. Exact multiracial categories not reported to uphold participant privacy and confidentiality. Age presented as *M* (*SD*), range. Other for full sample: Middle Eastern (*n*=1), Puerto Rican (*n*=3), Latine (*n*=5). Other for subsample: Puerto Rican (*n*=1). In full sample, caregiver *n* = 51. In sub-sample, caregiver *n* = 9.

**Table 3.** Participant Language, NVIQ, and Autism Traits.

| Variable | Full Sample (*N* = 73) |  |  | Subsample (*n* = 13) |  |  |
| --- | --- | --- | --- | --- | --- | --- |
|  | *M* | *SD* | range | *M* | *SD* | range |
| CELF-5 Receptive Language Index | 82.1 | 20.4 | 45-118 | 63.4 | 13.1 | 45-90 |
| CELF-5 Expressive Language Index | 77.4 | 18.8 | 45-116 | 65 | 19.2 | 45-91 |
| Peabody Picture Vocabulary Test-5<sup>th</sup> Ed. | 92.0 | 20.6 | 40-129 | 68.9 | 19.4 | 40-106 |
| Expressive Vocabulary Test-3<sup>rd</sup> Ed. | 94.0 | 17.7 | 49-128 | 74.3 | 15.7 | 49-106 |
| Syllable Repetition Task overall accuracy | 90.5 | 9.2 | 66-100 | 85.8 | 13.0 | 66-100 |
| Raven's 2 NVIQ | 90.7 | 16.1 | 47-135 | 83.4 | 13.9 | 62-106 |
| SRS-2 total *t*-score | 72.7 | 11.4 | 51-90 | 68.3 | 11.4 | 51-87 |
| BSCS overall sense of community | 3.1 | .9 | 1.4-5 | 3.1 | .7 | 2-4.3 |
| NLTS-2 unmet service needs | 3.3 | 3.2 | 0-14 | 2.6 | 2.3 | 0-8 |
| NLTS-2 barriers to services | 5.5 | 3.3 | 0-11 | 5.6 | 4.0 | 0-10 |

*Note.* CELF-5 = Clinical Evaluation of Language Fundamentals-5<sup>th</sup> Ed. (Wiig et al., 2013). Peabody Picture Vocabulary Test-5<sup>th</sup> Ed. (Dunn, 2019). Expressive Vocabulary Test-3<sup>rd</sup> Ed. (Williams, 2019). Syllable Repetition Task (Shriberg et al., 2009). Raven's 2 = Raven's 2 Progressive Matrices (Raven, 2018). SRS-2 = Social Responsiveness Scale-2<sup>nd</sup> Ed. (Constantino, 2012). NVIQ = nonverbal intelligence. Scores replaced using single imputation of variable means for: CELF-5, PPVT-5, EVT-3, SRT, and NVIQ scores (*n* = 1), as well as SRS-2 total *t*-scores (*n* = 2). Twenty-five participants were ages 22 or older. BSCS = Brief Sense of Community Scale, with a range of 0 to 5, or strongly disagree to agree (Peterson et al., 2008). NLTS-2 = National Longitudinal Transition Survey-2 (NLTS-2; Newman et al., 2011). Unmet service needs possible range = 0 to 16, and unmet barriers to services possible range = 0 to 12 (Newman et al., 2011; Taylor & Henninger et al., 2015).

### Measures

#### Sociodemographics

Per reporting guidelines (APA, 2019; Flanagin et al., 2021), autistic participants and caregivers provided race, ethnicity, sex assigned at birth, and gender for the autistic participants. Race and ethnicity used U.S. Census categories (Office of Management and Budget, 1997), with options to write in and to select multiple options.

#### Language Skills

Participants completed normed assessments of structural language (anonymized; Magiati et al., 2014); see Table 3. The aim of using normed assessments was to provide indices that align with service eligibility (Burke et al., 2023; Selin et al., 2022). Overall receptive-expressive language was assessed by the Clinical Evaluation Language Fundamentals-5^th^ Ed. (CELF-5; Wiig et al., 2013). For participants over age 21, age 21 norms were used per prior studies of adults ages 18 to 49 (Botting, 2020; Clegg et al., 2021; Fidler et al., 2011). Characterizing measures included receptive and expressive vocabulary, as assessed by the Peabody Picture Vocabulary Test-5^th^ Ed. (Dunn, 2019) and Expressive Vocabulary Test-3^rd^ Ed. (Williams, 2019), and nonword repetition, as assessed by the Syllable Repetition Task (Shriberg et al., 2009).

#### NVIQ

Nonverbal ability was assessed using the Raven’s Progressive Matrices-2^nd^ Ed. (ages 4-90; Raven et al., 2018). The Raven’s 2 does not use language and is untimed, which enhances accessibility (Grondhuis et al., 2018). Participants are presented with a visual stimulus and five pictures. In each stimulus, part of the picture is missing. Participants select one picture from the field of five to complete the picture.

#### Autism Traits

Autism traits were measured using the SRS-2, which has caregiver and self-report forms for youth and adults (Constantino, 2012). Respondents indicate the frequency of 65 items. Item-scores produce domain *t*-scores, a social communication impairment *t*-score, and an overall *t*-score. Total *t*-scores of ≤59 indicate sub-clinical, 60 to 65 mild, 66 to 76 moderate, and >76 high levels of autism traits.

#### Social Drivers of Health

Sense of community was measured using the Brief Sense of Community Scale, a validated scale for diverse youth and adults (Peterson et al., 2008). Respondents rate eight statements for agreement on a five-point scale, yielding an overall score (Peterson et al., 2008). Unmet service needs and barriers to services were measured using adapted items from the National Longitudinal Transition Study-2 (Newman et al., 2011), per prior work (Taylor & Henninger, 2015). Respondents reported if: a) each of 16 services were received (psychological, speech-language, vocational, aide, medical, occupational, tutor, transportation, social work, assistive technology, respite, reader/interpreter, physical therapy, mobility, audiology, other), b) each service is needed, and c) each of 12 items are barriers to meeting service needs (cost, location, doctor/specialist does not accept insurance, not available, scheduling conflicts, ineligible, lack of information, transportation, quality, lack of time, language barrier, physical accessibility). Item scores provide total counts.

#### Self-Determination

Self-determination was assessed using the SDI and a semi-structured interview. Participants ages 13 to 22 in secondary school completed the SDI:SR (Shogren et al., 2020). Those over age 18 and not in secondary school completed the SDI:AR (Shogren et al., 2021a). If caregivers were available, they completed the SDI:PTR (Shogren et al., 2021a). The interview followed the sequence of SDI items and used open-ended questions to understand nuances in responses; see [anonymized]. After discussing each item, respondents made a quantitative response (Shogren et al., 2015).

### Data Processing

Two trained research assistants independently scored and checked measures. Categorical cutoffs were NVIQ < 70 for intellectual disability (American Psychiatric Association, 2013), ≤ - 1.25 *SD* on at least two language measures for language impairment (anonymized; Tomblin et al., 1997), and SRS-2 total *t*-score > 76 for high levels of autism traits (Constantino, 2012). CELF-5 core language and Raven’s 2 standard scores were centered on *M* = 100. SRS-2 total *t*-scores were centered on 59, the subclinical threshold for autism traits. For the second aim, the first, second, fifth, and last authors transcribed the interviews and removed any identifiable information. Next, a trained research assistant who did not know study purpose or clinical status checked point-by-point accuracy at the level of utterance boundaries and words (Finestack et al., 2014). All disagreements were discussed until consensus was reached.

### Analysis

#### Statistical Analysis

Prior to analysis, data were checked for multicollinearity, homoscedasticity, normality, and missingness. Missing data were: (a) language and NVIQ scores (*n* = 1; did not complete assessment); (b) SRS-2 scores (*n* = 2; *n* = 1 missing form, *n* = 1 did not complete SRS-2); and (c) SDI self-report (*n*= 4 did not complete when other data available). Missing data were replaced using predictive mean matching with one imputation in SPSS 29 (IBM Corp., 2023; Little & Rubin, 2019). Analysis used an *a priori* significance level of .05. Independent sample *t*-tests showed nonsignificant differences by form (SDI:SR versus SDI:AR) in SDI self-report scores, *p* = .323, and associated SDI:PTR scores, *p* = 577. Thus, scores from SDI:SR and SDI:AR forms were combined into one group: SDI self-report. Associated SDI:PTR scores forms were also combined into one group: SDI:PTR.

Given small sample size, point-biserial correlations and Spearman correlations identified significant effects to use as predictors in regression (Spearman, 1904). Interpretation of effect sizes were *r* = 0.1 as small, 0.3 moderate, and 0.5 large (Cohen, 1988). Generalized linear models estimated the extent to which SDI scores and SDI:PTR scores were predicted from categorical individual differences (language impairment, intellectual disability, high levels of autism traits) and social drivers of health (sense of community, number of unmet service needs, number of unmet barriers). Analyses were repeated with general linear models to estimate effects of continuous individual differences (CELF-5 core language, NVIQ, and SRS-2 total *t*-scores). Models were fit using restricted maximum likelihood with *mixed* and GLM procedures in SPSS 29 (IBM Corp., 2023), with fixed effects of predictors and by-participant random intercepts.

#### Thematic Analysis

For the second aim, primary outcomes were organizing interview themes. All authors used thematic analysis to identify and organize themes (Braun & Clarke, 2012). Authors familiarized themselves with the data, and then the first three authors inspected the data to generate preliminary codes (Braun & Clarke, 2012). Next, coders met to discuss codes, iteratively refine preliminary codes, and to develop a codebook, with definitions and example quotes (Braun & Clarke, 2012). Then, coders organized codes into networks of themes, with organizing themes and sub-themes (Braun & Clarke, 2012). The full research team reflected on, reviewed, and revised the codes and themes. All stages of analysis were iterative and introspective, with no stopping point other than saturation (Braun & Clarke, 2012). To protect participant privacy and confidentiality, analysis considered themes by respondent group versus specific dyads (O’Brien et al., 2014).

## Results

### Role of Individual Differences and Social Drivers of Health in SDI Scores

#### Categorical Approach

Point-biserial correlations revealed significant, small to moderate effects between SDI:PTR scores and high levels of autism traits, sense of community, and number of unmet service needs; see Table 4. Generalized linear models examined predictive effects of SDI:PTR scores using a gamma link identity function; see Table 5. In the empty model, SDI:PTR scores significantly varied, τ^2^ = 522.32, *z* = 5.05, *p* = < .0001. When including fixed effects, model fit improved. AIC and BIC were smaller, and marginal pseudo-*R*^2^ increased to .407, indicating variance explained for by fixed effects. Intercept variance remained significant, τ^2^ = 321.20, *z* = 4.90, *p* = < .0001. Given sense of community and unmet service needs, high levels of autism traits were associated with a 15.96 decrease in SDI:PTR scores. Given high levels of autism traits and unmet service needs, for every unit increase in overall sense of community, SDI:PTR scores were expected to be higher by 11.09. Overall, categorical approaches to individual differences had limited predictive value for SDI scores.

**Table 4.**
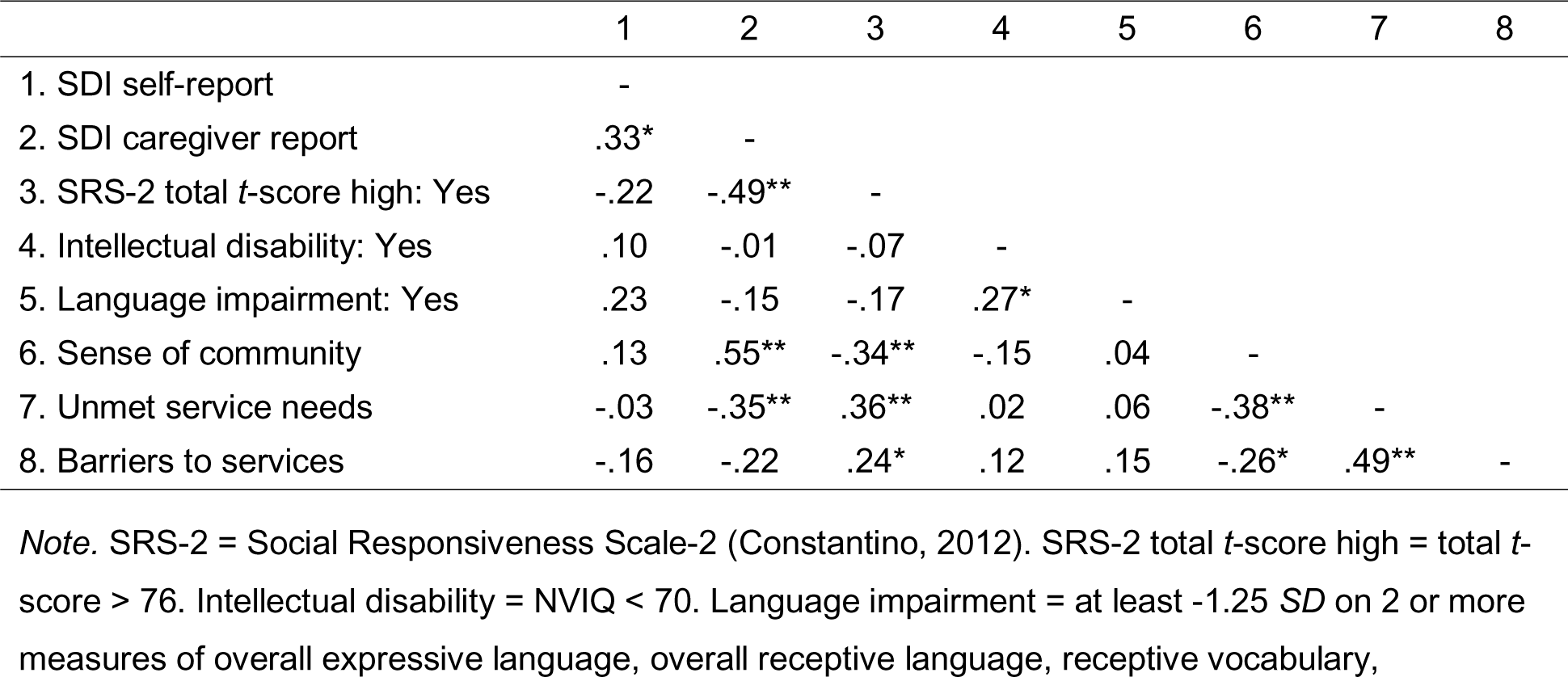
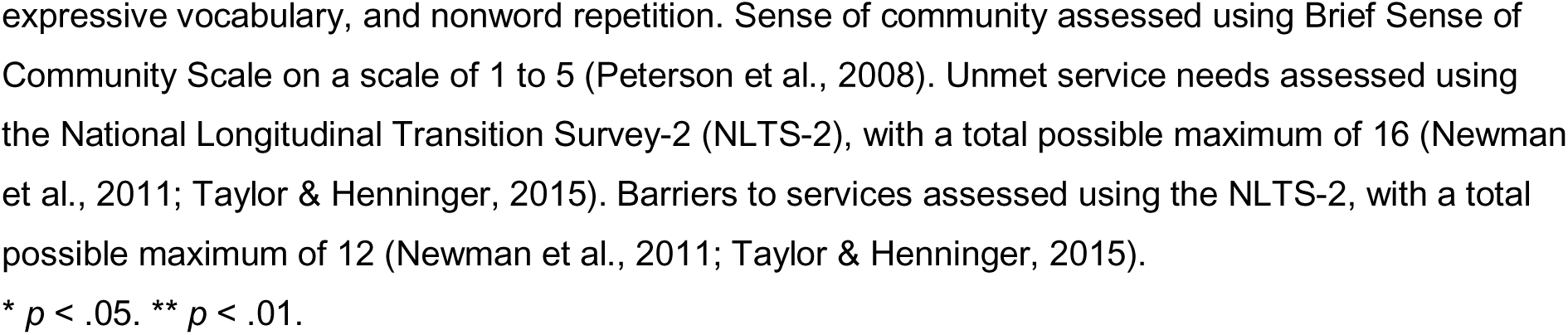
Point Biserial Correlation Coefficients of SDI Scores, Language Impairment, Intellectual Disability, High Levels of Autism Traits & Social Drivers of Health.

**Table 5.** Generalized Linear Mixed Model Results of Using Categorical Individual Differences and Social Drivers of Health Measures to Predict SDI Caregiver Scores.

| Parameter | Model 1 |  |  | Model 2 |  |  |
| --- | --- | --- | --- | --- | --- | --- |
| | $\beta$ | SE | 95% CI | $\beta$ | SE | 95% CI |
| Intercept | 59.62*** | 3.17 | 53.25, 65.98 | 15.22 | 11.71 | -8.32, 38.75 |
| SRS-2 total t-score: high |  |  |  | -15.96** | 5.70 | 4.49, 27.43 |
| BSCS overall sense of community |  |  |  | 11.09** | 3.18 | 4.70, 17.48 |
| Unmet service needs |  |  |  | -.66 | .91 | -2.49, 1.17 |
| *Pseudo-R<sup>2</sup> marginal* | .000 |  |  | .407 |  |  |
| *Pseudo-R<sup>2</sup> conditional* | 1.00 |  |  | 1.000 |  |  |
| *AIC* | 472.105 |  |  | 433.447 |  |  |
| *BIC* | 475.718 |  |  | 436.922 |  |  |
| *Adjusted ICC* | 1.00 |  |  | 1.000 |  |  |
| *Conditional ICC* | 1.00 |  |  | .593 |  |  |

*Note.* Model presented as $\beta$ (SE) [95% CI]. = $\beta$ estimates of fixed effects. SE = standard error. 95% CI = 95% confidence interval. AIC = Akaike's Information Criterion. BIC = Schwarz's Bayesian Criterion. CELF-5 and SRS-2 estimates based on centered predictors. By-participant random intercepts were included in the models. SRS-2 total $t$ -score: high = Social Responsiveness Scale-2nd Ed. total $t$ -score > 76 (Constantino, 2012). BSCS = Brief Sense of Community Scale (Peterson et al., 2008), with a possible range of 1 to 5. Unmet service needs = number of unmet service needs, as assessed by adapted items from the National Longitudinal Transition Survey-2 (Newman et al., 2011; Taylor & Henninger, 2015), with a total possible of 16.
\*= $p < .05$ , \*\* $p < .01$ , \*\*\* $p < .001$ .

#### Continuous Approach

Spearman’s rank-order correlations revealed significant associations between SDI self-report scores with SRS-2 total *t*-scores and CELF-5 core language scores; see Table 6. SDI:PTR scores significantly associated with SRS-2 total *t*-scores, sense of community, and unmet service needs. Separate general linear models examined predictive effects on SDI self-report and SDI:PTR scores. In the empty model, SDI self-report scores significantly varied, τ^2^ = 105.64, Wald *z* = 3.04, *p* = .001. Including fixed effects improved model fit; see Table 7. AIC and BIC were smaller, and marginal and conditional pseudo-*R*^2^ increased. Intercept variance remained significant, τ^2^ = 90.14, Wald *z* = 3.08, *p* = .001. Given SRS-2 total *t*-scores, for every point increase in CELF-5 scores, SDI self-report scores were expected to be lower by .17 relative to the intercept (the expected SDI self-report score for a person with a CELF-5 core language score of 100 and SRS-2 total *t*-score of 59). Given CELF-5 core language scores, for every point increase in SRS-2 total *t*-scores, SDI self-report scores were expected to be lower by -.42 relative to the intercept.

**Table 6.**
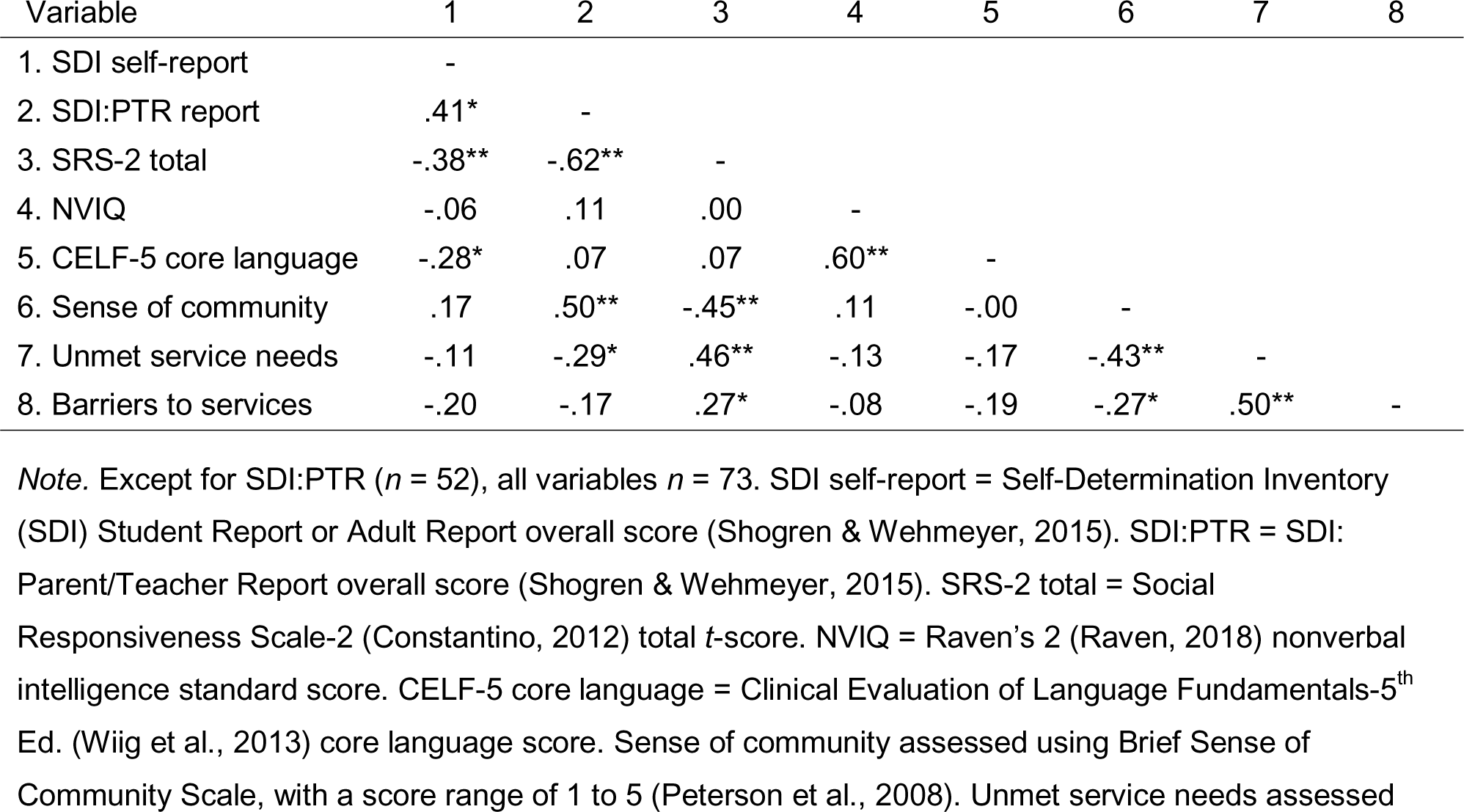
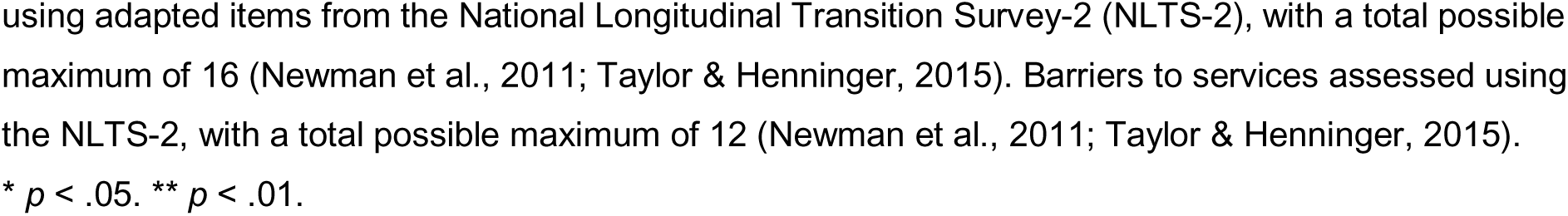
Spearman’s Correlations Coefficients of SDI Scores, Individual Difference Measures, and Social Drivers of Health.

**Table 7.**
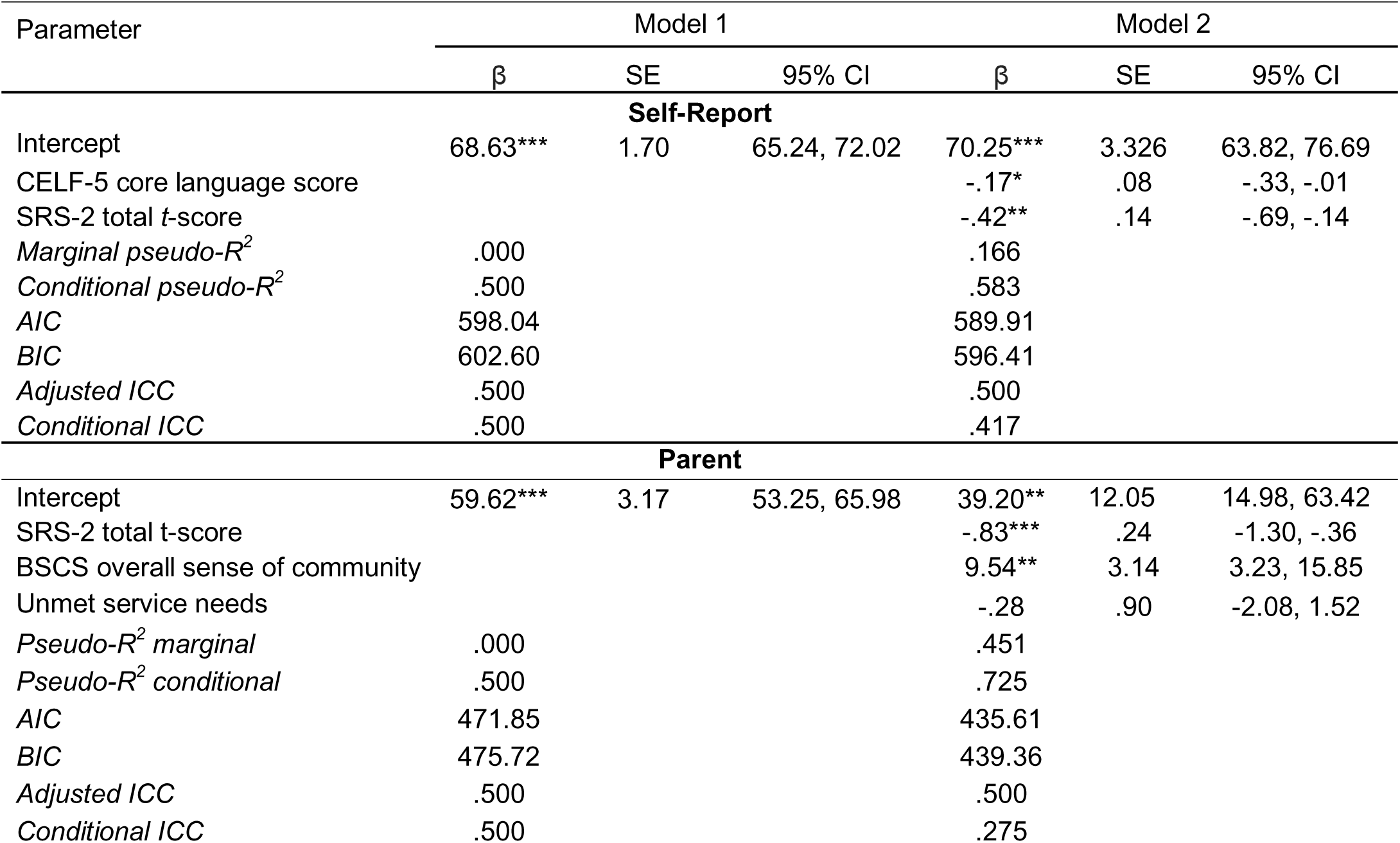
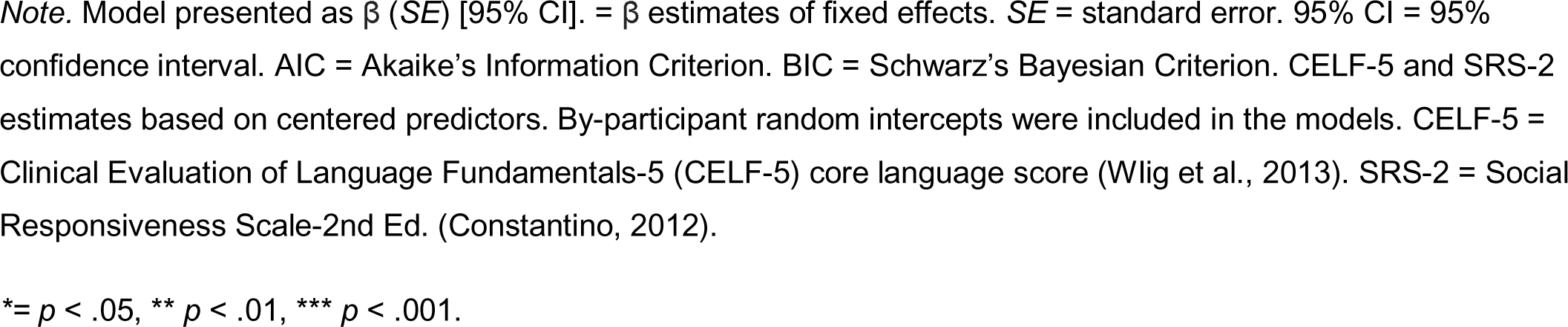
Mixed-Effects Model Results of SDI Self-Report and SDI Parent Report Scores.

The empty model showed significant variation in SDI:PTR scores, τ^2^ = 261.16, Wald *z* = 2.57, *p* = .005. When including fixed effects, model fit improved; see Table 7. Intercept variance remained significant, τ^2^ = 148.18, Wald *z* = 2.65, *p* = .004. Covarying for sense of community and unmet service needs, for every one point increase in SRS-2 total *t*-scores, SDI:PTR scores were expected to be lower by -.83 relative to the intercept. Controlling for SRS-2 total *t*-scores and unmet service needs, for every unit increase in sense of community, SDI:PTR scores were expected to be higher by 9.54 relative to the intercept.

#### Summary

Predictors of SDI:PTR overall scores were similar when using categorical or continuous approaches to individual differences. However, SDI self-report scores were only predicted by continuous approaches. One question is how qualitative data compare to SDI scores.

### Qualitative Themes from the SDI

Three organizing themes emerged from interviews from autistic participants with language impairment that aligned with Causal Agency Theory (Shogren & Wehmeyer, 2015). Consistent with DisCrit and Diversity Science, themes revealed both nuanced experiences of marginalization (Annamma et al., 2013) and heterogeneity of experiences (Plaut, 2010).

#### Contextual Factors in Beliefs about Setting and Reaching Goals

Given a social-ecological model of disability (Bronfenbrenner, 1977), context reflects both individual differences and external reactions to individual differences. These interactions shape experiences of self-determination (Scott et al., 2021; Taylor et al., 2023). A critical part of context was the process by which environmental fit constrained beliefs about goal setting and attainment. For participants, context involved access to supports. For caregivers, context also included balancing supporting their family member’s goals with ensuring their safety.

##### Environmental Threats on Autonomy

Environmental threats to autonomy refer to constraints arising from where individuals live on beliefs about their ability to set and achieve goals (Chang et al., 2017). One participant shared his goals; see top of Subtheme 1 in Table 8. He lived in a community where environmental demands were high for anyone – in particular, for a Black autistic man with language impairment post-George Floyd. His goals did not pertain to actually learning how to use a phone. Instead, goals referred to having access to the means to navigate complex environmental demands, such as knowing how and when to call for help in the case of physical threat. He later explained his efforts to navigate these demands; see bottom of Subtheme 1 in Table 8. His caregiver explained they taught him to learn when to ask for help and to ask for help versus problem solve, given safety concerns; see Table 9.

**Table 8.**
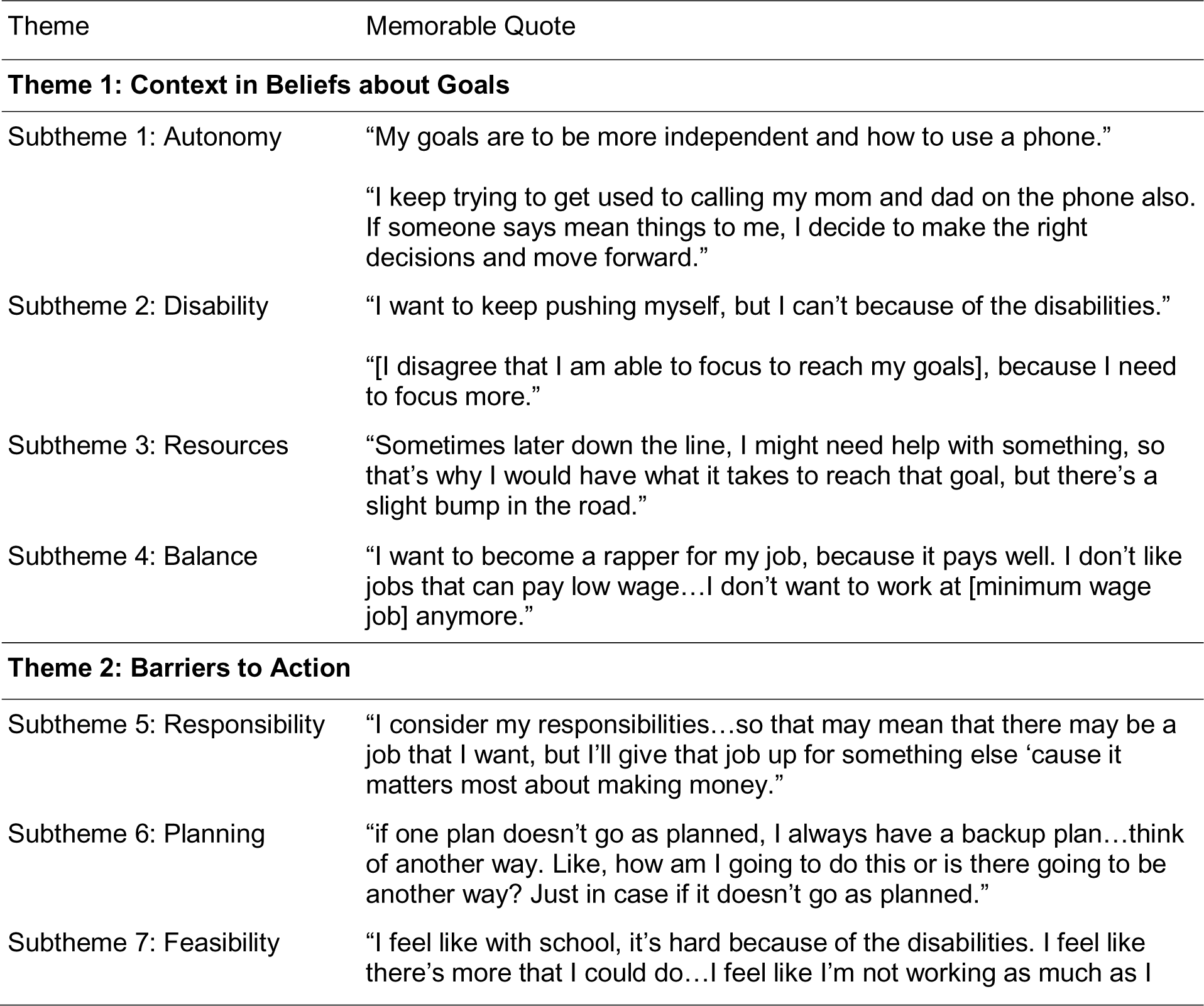
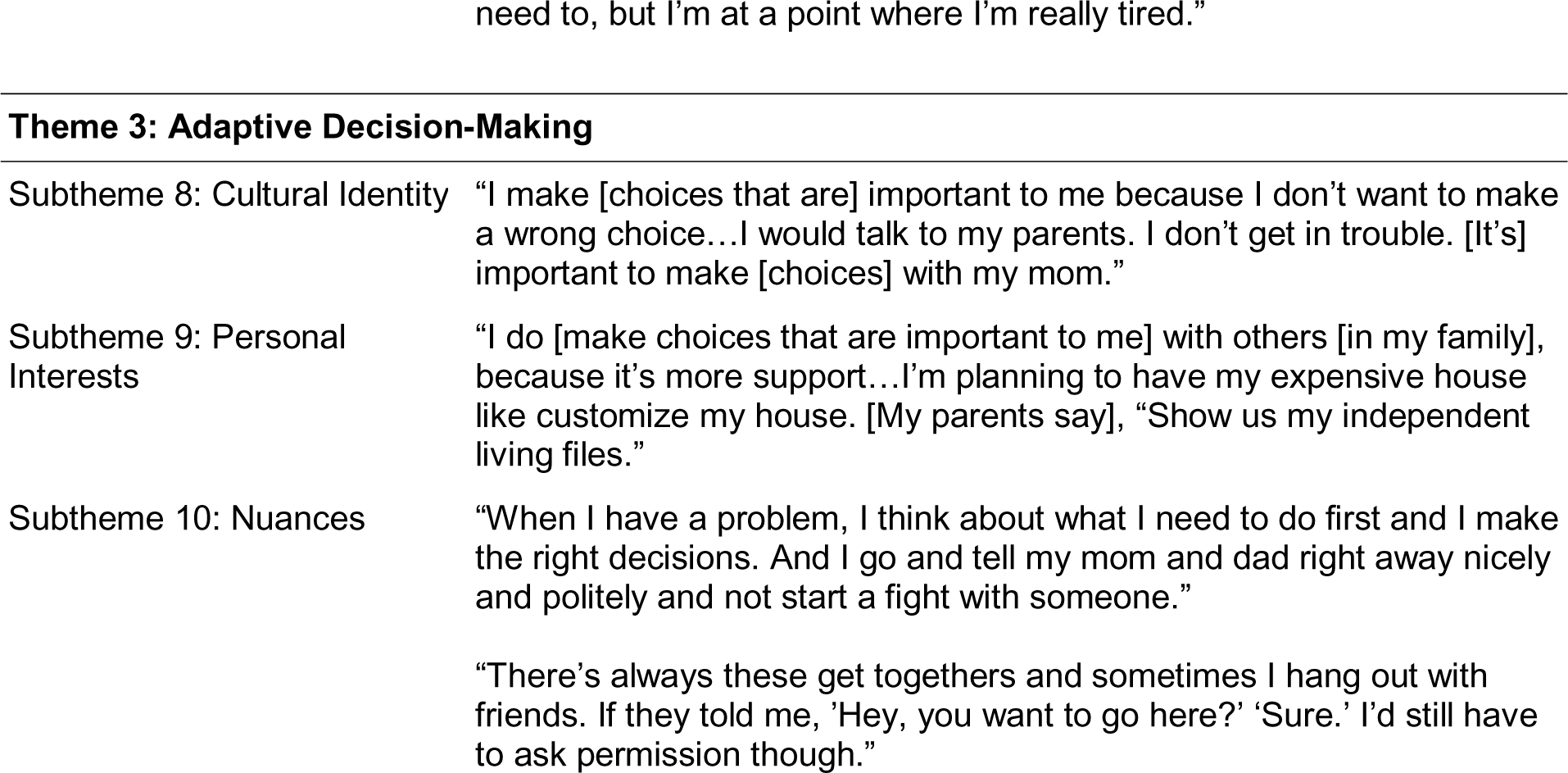
Themes and Memorable Quotes from Self-Determination Inventory Self-Report Interviews.

**Table 9.**
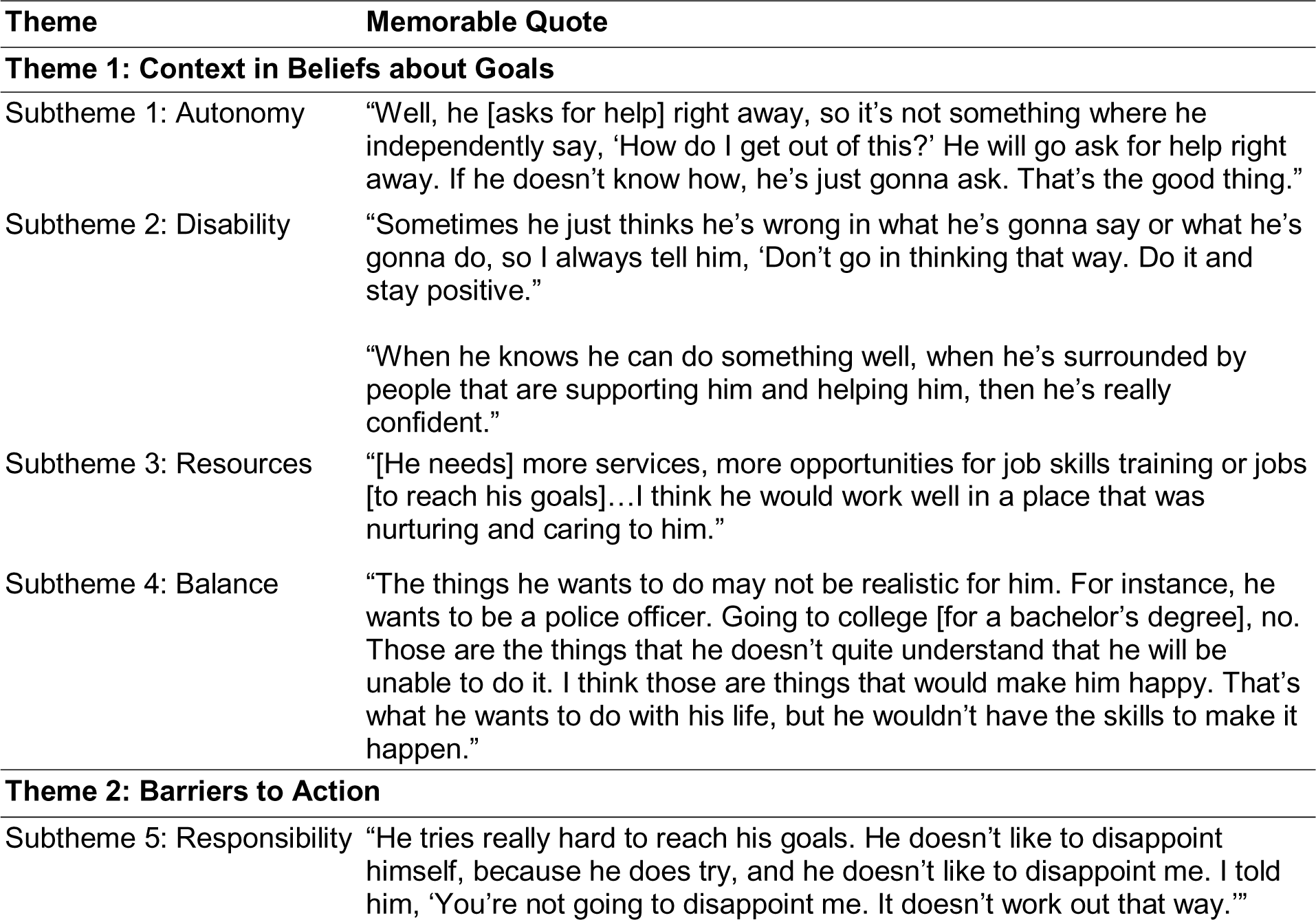
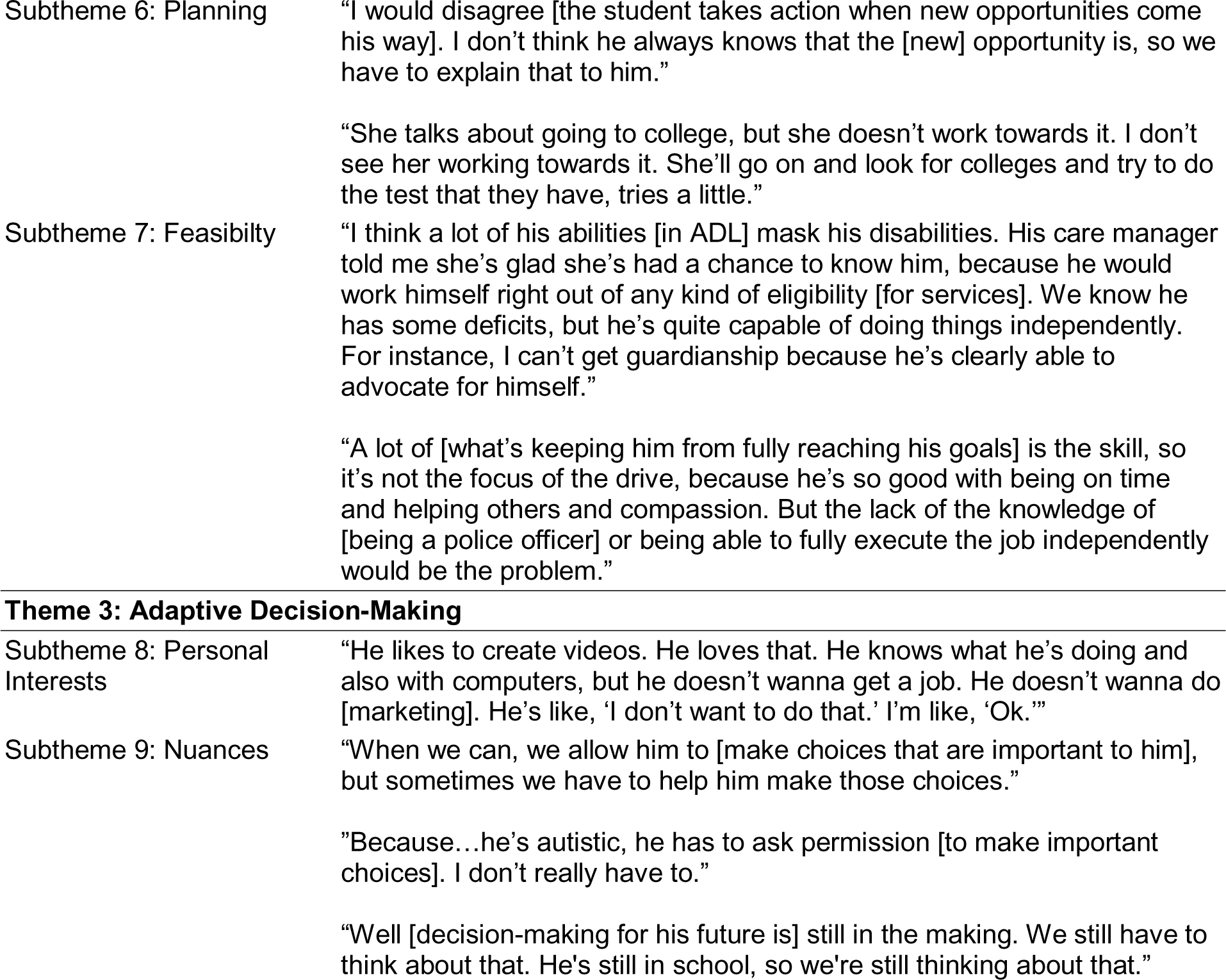
Themes and Memorable Quotes from Self-Determination Inventory Caregiver Interviews.

| Theme | Memorable Quote |
| --- | --- |
| **Theme 1: Context in Beliefs about Goals** |  |
| Subtheme 1: Autonomy | "Well, he [asks for help] right away, so it's not something where he independently say, 'How do I get out of this?' He will go ask for help right away. If he doesn't know how, he's just gonna ask. That's the good thing." |
| Subtheme 2: Disability | "Sometimes he just thinks he's wrong in what he's gonna say or what he's gonna do, so I always tell him, 'Don't go in thinking that way. Do it and stay positive.'"<br>"When he knows he can do something well, when he's surrounded by people that are supporting him and helping him, then he's really confident." |
| Subtheme 3: Resources | "[He needs] more services, more opportunities for job skills training or jobs [to reach his goals]...I think he would work well in a place that was nurturing and caring to him." |
| Subtheme 4: Balance | "The things he wants to do may not be realistic for him. For instance, he wants to be a police officer. Going to college [for a bachelor's degree], no. Those are the things that he doesn't quite understand that he will be unable to do it. I think those are things that would make him happy. That's what he wants to do with his life, but he wouldn't have the skills to make it happen." |
| **Theme 2: Barriers to Action** |  |
| Subtheme 5: Responsibility | "He tries really hard to reach his goals. He doesn't like to disappoint himself, because he does try, and he doesn't like to disappoint me. I told him, 'You're not going to disappoint me. It doesn't work out that way.'" |
| Subtheme 6: Planning | "I would disagree [the student takes action when new opportunities come his way]. I don't think he always knows that the [new] opportunity is, so we have to explain that to him." |
|  | "She talks about going to college, but she doesn't work towards it. I don't see her working towards it. She'll go on and look for colleges and try to do the test that they have, tries a little." |
| Subtheme 7: Feasibility | "I think a lot of his abilities [in ADL] mask his disabilities. His care manager told me she's glad she's had a chance to know him, because he would work himself right out of any kind of eligibility [for services]. We know he has some deficits, but he's quite capable of doing things independently. For instance, I can't get guardianship because he's clearly able to advocate for himself." |
|  | "A lot of [what's keeping him from fully reaching his goals] is the skill, so it's not the focus of the drive, because he's so good with being on time and helping others and compassion. But the lack of the knowledge of [being a police officer] or being able to fully execute the job independently would be the problem." |
| **Theme 3: Adaptive Decision-Making** |  |
| Subtheme 8: Personal Interests | "He likes to create videos. He loves that. He knows what he's doing and also with computers, but he doesn't wanna get a job. He doesn't wanna do [marketing]. He's like, 'I don't want to do that.' I'm like, 'Ok.'" |
| Subtheme 9: Nuances | "When we can, we allow him to [make choices that are important to him], but sometimes we have to help him make those choices." |
|  | "Because...he's autistic, he has to ask permission [to make important choices]. I don't really have to." |
|  | "Well [decision-making for his future is] still in the making. We still have to think about that. He's still in school, so we're still thinking about that." |

##### Impact of Experiences with Disability on Action-Control Beliefs

Experiences with disability impacted beliefs about agency in setting and reaching goals; see Subtheme 2 in Table 8. One participant commented on a need to focus. His caregiver explained that he is self-conscious about his limitations in communication relative to social norms, such as ease of using spoken language relative to community expectations for verbal fluency. Other caregivers also noted limitations in the self-efficacy of their family members. Specifically, participants were aware of their disabilities and felt less capable to set goals given environmental demands; see Subtheme 2 in Table 9.

##### Importance of Resources

Both groups spoke about the importance of supports to set and achieve goals. Consistent with autonomy, participants prioritized resources versus independence to achieve goals; see Subtheme 3 in Table 8. A caregiver noted the importance of supports in enhancing their child’s beliefs about their ability to set goals, which largely included employment; see Subtheme 3 in Table 9. Access has implications for considering environmental responsivity (or lack thereof).

##### Balancing Beliefs about Goal Setting with Feasibility

Participants emphasized the importance of feeling they could reach their goals while maintaining their sense of dignity. For example, one wanted to earn a wage commensurate with his sense of self-worth that was sufficient for buying a home; see Subtheme 4 in Table 8. Caregivers believed in the importance of supporting their family member in goal setting, but the feasibility of achieving goals was a major concern. This is especially salient when considering multiple marginalization (Annamma et al., 2013), as caregivers may not be able to ensure the safety of their family member in society; see Subtheme 4 in Table 9.

#### Barriers to Action

Barriers to action refers to material and immaterial personal and environmental factors that impact individual ability to take action to work toward goals. Responses reflected tension between participants working to achieve goals as self-directed agents and recognizing challenges. Understanding systemic discrimination on the experiences of minoritized autistic adolescents and adults helped frame responses in terms of actions and barriers (Crenshaw, 1989).

##### Sense of Responsibility

Participants expressed they wanted to act responsibly as adults but encountered challenges; see Subtheme 5 in Table 8. One shared his family had reduced access to opportunities over generations due to the legacy of racism (Crenshaw, 1989; Powell, 2012). Thus, despite having a career interest that aligned with his access needs, he did not want to perpetuate the cycle of poverty. In this case, environmental barriers pertained to both disability and race. In turn, a caregiver commented on their son’s work ethic; see Subtheme 5 in Table 9. Follow-up indicated he held himself to a standard of success on the first try, based on observations of nonautistic age peers. Even with family telling him otherwise, he believed any failure in taking action to reach their goals was due to his limitations and not social norms.

##### Strategic Planning

Both groups spoke about flexibility in planning to take action to reach short- and long-term goals, challenging theories of autism (American Psychiatric Association, 2013); see Subtheme 6 in Table 8. One participant explained he had previously been unable to tolerate change, such as public transportation delays, or activities that did not align with his personal interests, resulting in multiple meltdowns per week. After a caregiver passed away during the pandemic, becoming more flexible was important. Planning sometimes required supports; see top of Subtheme 6 in Table 9. In this scenario, navigating post-secondary programming was inaccessible to their child. Thus, the caregiver translated information into accessible visuals and text to support their child taking action. Inaccessibility as a barrier to action emerged elsewhere. One caregiver spoke about gaps between their child’s actions and goal of attending a STEM degree program due to inaccessibility of entrance exams; see bottom of Subtheme 6 in Table 9.

##### Balancing Action with Feasibility

Respondents spoke about juggling goal-oriented action with disabilities. One participant had a strong interest in school but limited capacity to act due to the inaccessibility of navigating social interactions, language demands, and services; see Subtheme 7 in Table 8. Caregivers often spoke of their children’s persistence; see top of Subtheme 7 in Table 9. One might believe that the participant does not need supports unless considering environmental demands for Black autistic adults with language impairment and intellectual disability. Caregivers shared stories of others taking advantage of their children (e.g., cheating them when buying items) or not realizing their children had disabilities (e.g., misinterpreting actions as aggression, leading to harm), especially if their children were physically adults; see bottom of Subtheme 7 in Table 9.

#### Adaptive Decision-Making

The third theme involved integrating different factors to make decisions. Decision-making was a dynamic process, with influences from culture and personal interests (Plaut, 2010), as well as inadequate environmental supports (Annamma et al., 2013). Safety in the environment associated with racism and ableism was a concern.

##### Cultural Sense of Identity

Cultural sense of identity pertained to how respondents conceptualizing themselves. Per prior work, respondents shared that autonomy involved the family as a unit versus the individual (Leake & Boone, 2007; Trainor, 2005). Yet, cultural norms and experiences of disability did not exist in a vacuum; see Subtheme 8 in Table 8. The response might not appear to reflect cultural norms, because the participant mentioned “getting in trouble.” In actuality, “getting in trouble” reflected: 1) being able to navigate environmental demands, such that the participant *had* gotten in trouble in the past when making independent choices (e.g., getting lost when taking public transportation alone without knowledge or skills of how to navigate public transportation); and 2) being of a cultural background where the decision-making unit was the family.

##### Personal Interests

Participants often drew from their personal interests to make decisions. This perspective was not mutually exclusive with cultural norms. For instance, one participant perceived making decisions autonomously *and* actively consulting family members in an extended, multi-generational household; see Subtheme 9 in Table 8. Understanding that there are different cultural notions of autonomy and family structure is important for appreciating personal interests. Elsewhere, a caregiver spoke about how their child had a personal interest and skills, which she and her family supported. Yet, translating their interests into decision-making was challenging; see Subtheme 8 in Table 9.

##### Nuanced Autonomy in Decision-Making

Decision-making often involved responding to environmental demands. Participants reported making decisions with family, but not because of lack of autonomy; see Subtheme 10 in Table 8. In the former case, decision-making involved active reflection and turning to family for support if he encountered a problem. In the latter case, decision-making required asking a caregiver for permission to go out with friends for safety concerns. These examples suggest the impact of environmental demands for minoritized autistic adults with language impairment on autonomy. Caregivers echoed this theme; see Subtheme 9 in Table 9. Prior experience with independent decision-making led to one participant nearly providing home address and financial information to strangers online and another experiencing threats to physical safety. Per DisCrit (Annamma et al., 2013), even with support of self-determination, minoritized families must grapple with the impossibility of ensuring safety given numerous incidents involving race and disability. Overall, cultural, personal, and environmental factors made decision-making a dynamic process; see bottom of Subtheme 9 in Table 9.

#### Summary

Interview themes from autistic individuals with language impairment and caregivers showed the importance of context in self-determination. Responses reflected both individual experiences and environmental factors.

## Discussion

Findings supported the relevance of categorical versus dimensional approaches in predicting self-determination (Kover & Abbeduto, 2023). This study also extends prior work documenting complex intersectional effects of race and disability on self-determination (Scott et al., 2021; Taylor et al., 2023). However, key differences exist.

### Influences in Self-Determination: Measurement Matters

In this study, SDI self-report scores were similar to SDI:SR latent means in diverse groups of autistic students (67.4 versus 70.58 to 72.99; Shogren et al., 2018a). However, SDI:AR scores were lower than in prior work (71.1 versus 80.45; Hagiwara et al., 2021). Age might explain this difference. Hagiwara et al. (2021b) found SDI:AR estimates increased each year in adults up to age 71, with increased opportunities for autonomy in adulthood. There were also differences in multi-informant concordance. Here, concordance of self- and caregiver-reported scores was higher (*r* = .34) than in prior work comparing self- and teacher-reported scores (*r* = .07; Shogren et al., 2021a). It could be that teachers interact with students in more limited contexts than caregivers or have different perspectives on self-determination due to misalignment, as teachers of minoritized students are primarily white (Shogren et al., 2021a).

A second point is that predictors of self-determination varied by conceptualization of individual differences. High levels of autism traits predicted SDI:PTR overall scores, while primary disability label did not predict self-determination in adults (Hagiwara et al., 2021a). To align with real-world variability, this study did not use primary disability label; it is unknown if a high level of autism traits yields a clinically meaningful difference in self-determination (Chatham et al., 2018). Continuous approaches had different effects. Language scores predicted SDI self-report scores, while autism traits predicted both SDI self-report and SDI:PTR scores. On one hand, language has predicted outcomes in autism, while effects of autism traits are more inconsistent (Magiati et al., 2014). Given its importance in postsecondary outcomes (Shogren et al., 2015, 2017a), self-determination might moderate the relationship between autism traits and outcomes. Moreover, using a language-based modality may have influenced outcomes.

Effects of social drivers of health only partly aligned with expected results. As expected (Cheak-Zamora et al., 2020), unmet service needs did not predict SDI:PTR scores. However, unmet service needs did not predict SDI self-report scores, nor did barriers to services predict SDI self-report or SDI:PTR scores (Ishler et al., 2023; Taylor & Henninger, 2015). Better assessments of service needs and barriers may be needed (Burke et al., 2023). Similarly, sense of community predicted SDI:PTR, but not SDI self-report scores. While the effect on SDI:PTR scores was expected (Daley et al., 2018), the lack of effect for SDI self-report was not. Ultimately, findings motivate attention and care in assessment approach.

### Implications for Autistic Individuals

One question is what themes mean for minoritized autistic individuals. Themes pertained to contextual factors in beliefs, barriers to action, and decision-making, differing from findings from Black youth with developmental disabilities (e.g., historical undertones, spatialization of racialization, proxies for racial bias, interest convergence; Taylor et al., 2023). Both studies used DisCrit (Annamma et al., 2013) as a grounding theory, but studies differed. We explored self-determination in minoritized autistic adolescents and adults versus transition and self-determination in Black youth (Taylor et al., 2023). One commonality across studies, however, was the role of the environment in shaping experiences at the intersection of race and disability (e.g., environmental threats to autonomy). These themes have implications for characterizing the transition to adulthood.

Given the importance of sense of community, targeting environmental fit must eradicate systemic bias (Lai et al., 2020). Removing barriers, such as inadequate access to supports, could reduce misfit by embracing each person as they are. Commensurate with patterns in autism research (Anderson et al., 2018b; Howlin & Taylor, 2015), findings call for attention to how social drivers of health shape the transition to adulthood. Interviews revealed numerous strengths in participants with language impairment. As over 50% of the entire sample had language impairment and approximately 30% had NVIQ < 70 to 84, determining how to further merge individual differences with strengths and systemic factors requires meaningfully partnering with minoritized autistic individuals (Maye et al., 2021).

### Limitations

This study had several limitations. Sampling did not extend to individuals who are minimally speaking (Koegel et al., 2020). Examination of environmental factors was limited, amid a lack of best practices for assessing social drivers of health (Anderson et al., 2018b; Girolamo et al., 2023d). For instance, household income and other indicators may not be meaningful without context (e.g., income relative to cost of living; APA, 2020; JAMA Network Editors, 2020). Here, determining how to report social drivers of health and sociodemographics while maintaining privacy and confidentiality led to not including comprehensive sociodemographic information [anonymized].

### Future Directions

Limitations also offer pathways forward. For replicability, larger sample analysis leveraging theory-driven approaches, such as the ones in this study, is needed. In addition, larger sample analysis will allow for understanding the directionality of relationships between self-determination and predictors of environmental fit (Lai et al., 2020). Themes were consistent with prior research, such as community participation (Kim, 2019). A next step is understanding how SDI scores relate to health outcomes. Compared to extensive interviews, SDI scores might be easier to adopt in clinical settings. Last, understanding the relationship between language and self-determination over time is important (Hagiwara et al., 2021a), both for developing supports and understanding development.

### Conclusion

Assessing self-determination yielded new information in minoritized autistic adolescents and young adults with wide-ranging language and cognitive profiles and their caregivers. Findings pointed to the importance of individual differences and sense of community. While self-determination encompasses environmental demands, data make clear the importance of conceptualizing personal, family, and cultural factors together with social drivers of health.

## Data Availability

All data are not available due to participants opting to not share their data in a dynamic consent process. Information on data structure is available upon reasonable request to the authors.

## Acknowledgements

TG was supported by an American Speech-Language-Hearing Foundation New Investigators Research Grant (PI: Girolamo) and NIH L70DC021323. RC was supported by NIH T32DC017703 (PD: Eigsti).

